# Stimulation-evoked orbitofrontal activity as a biomarker for DBS personalization in depression

**DOI:** 10.64898/2026.06.25.26356607

**Authors:** Layth S. Mattar, Lily Chamakura, Amirreza Alijanpourotaghsara, Sameer Rajesh, Atefeh Ghazavi, Evangelia Tsolaki, Victoria Gates, Anusha Allawala, Nicole R. Provenza, Kala Bailey, Sanjay Mathew, Denise Oswalt, Garrett P. Banks, Wayne K. Goodman, Sameer A. Sheth, Sarah R. Heilbronner, Nader Pouratian, Eleonora Bartoli

## Abstract

Finding suitable therapies for treatment-refractory neuropsychiatric disorders constitutes a major goal for translational neuroscience. Deep brain stimulation shows promise for treatment resistant depression, but treatment efficacy varies substantially across patients. Objective, electrophysiologically driven strategies to optimize deep brain stimulation for treatment resistant depression could greatly improve clinical efficacy by minimizing the trial-and-error approach needed to personalize stimulation settings. This may not only reduce the delay between the start of the treatment and symptom improvement, but also enable acute, real-time verification of circuit engagement, advancing our understanding of the mechanism mediating antidepressant effects.

Here, we investigate whether cerebro-cerebral evoked potentials elicited through different deep brain stimulation configurations could be used to guide stimulation personalization for treatment resistant depression. Cerebro-cerebral evoked potentials offer a fast, objective way to identify regions engaged by stimulation, revealing the effective connectivity pattern of the stimulated location. Data were collected from eight patients with treatment resistant depression who received dual bilateral deep brain stimulation devices targeting the subcallosal cingulate and ventral capsule/ventral striatum. During an initial in-hospital monitoring period, single-pulse electrical stimulation was delivered through the deep brain stimulation devices and cerebro-cerebral evoked potentials were recorded through temporary stereo-electroencephalography probes across fronto-temporal regions. Patients underwent several outpatient stimulation programming sessions over the course of 9 months to identify the stimulation configurations leading to the greatest improvement in depressive symptoms. We retrospectively analysed cerebro-cerebral evoked potentials obtained in response to stimulation of different stimulation configurations to identify features distinguishing the clinically effective configurations.

The deep brain stimulation configurations leading to the greatest improvement in depressive symptoms were associated with significantly larger evoked potentials in the orbitofrontal cortex and showed an increased number of evoked potentials across dorsal and ventral prefrontal regions. Waveform similarity analysis revealed a gradient in therapeutic effects, such that multiple alternative stimulation configurations led to similar symptom improvement. The vast deep brain stimulation parameter space might contain a configuration subspace defined by comparable therapeutic effects. In addition, evoked potentials obtained from single-pulse and from bursts of high-frequency stimulation displayed similar spatial patterns, suggesting that either method might be able to identify the configuration best engaging the circuit mediating the clinical response.

Together, these findings provide proof-of-principle evidence that stimulation-evoked prefrontal responses reflect network engagement associated with antidepressant effects. Cerebro-cerebral evoked potentials may offer an objective and acute strategy to guide contact selection in deep brain stimulation for treatment resistant depression.

## Introduction

Major depressive disorder, a condition which can present with symptoms such as persistently low mood, anhedonia, lethargy, sleep disturbances, suicidal thoughts and more, has been projected to become the largest cause of burden of disease worldwide by 2030^1–3^. Neurally, depression is not due to the abnormality of a single brain region in isolation, but due to discord within and between multiple networks^4,5^. Many of those experiencing major depression can achieve remission through singular or combined conventional therapeutics like psychotherapy and pharmacotherapy, but about 30% of diagnosed individuals fail to respond to these traditional treatment options^6,7^, a condition often termed treatment resistant depression (TRD)^8^. While therapeutic options for TRD remain limited, experimental deep brain stimulation (DBS) of the subcallosal cingulate (SCC) or ventral capsule/ventral striatum (VC/VS) has shown promising results for this patient population^9–11^. However, not all patients benefit from this line of treatment, and currently we lack an understanding of the factors driving this variability in therapeutic response as well as strategies to increase its efficacy.

One factor contributing to outcome variability is that DBS use and mechanism of action are less established for treatment resistant psychiatric disorders and their heterogeneous neural correlates^12–15^. SCC DBS likely improves depression symptoms by modulating and reducing hyperactivity in the affective, salience and default mode networks^16–19^. Physiological changes caused by stimulation of the VC/VS in TRD patients remain less clearly characterized, but modulations of activity in the reward circuit have been reported across various studies^20–23^. Although SCC and VC/VS seem to engage distinct networks, one study demonstrated that stimulation of these targets generates widespread evoked potentials in the ventral prefrontal cortex^21^, potentially implicating this region in shared mechanisms of response. Even without a full mechanistic understanding of the therapeutic effects, a variety of methods are currently employed to guide DBS and improve its efficacy.

One method for the optimization and personalization of the DBS device placement relies on diffusion imaging combined with biophysical modelling to maximize the engagement of white matter tracts, with demonstrated improvements in antidepressant effects^24–27^. However, these indirect methods do not allow acute, real-time verification of stimulation-driven circuit engagement. By contrast, objective biomarkers from electrophysiological monitoring^28,29^, such as intracranial or scalp-electroencephalography evoked potentials in response to stimulation, could provide complementary information with respect to imaging and modelling approaches^30^. This approach is gaining traction in DBS for movement disorders, such as Parkinson’s and essential tremor, based on mounting evidence that stimulation-evoked resonant neural activity (ERNA) in the basal ganglia predicts clinical improvement^31–33^. Detecting and monitoring a similar objective, acute biomarker of circuit engagement in DBS for TRD would be particularly impactful given that symptom improvement can take weeks to months to become clinically meaningful^34^. This would accelerate the ability to identify the most promising contacts on the DBS lead, effectively reducing trial-and-error attempts to optimize the stimulation configuration (i.e., combination of contacts) for each patient. To date, we do not know whether an objective, acute stimulation-evoked biomarker capturing engagement of the circuit mediating antidepressant effects exists, nor where to look for it across the complex mixture of networks implicated in TRD. There is preliminary evidence of an acute stimulation-evoked pattern in TRD patients that can be measured non-invasively^35^, but its anatomical origin and relation to antidepressant effects have not been established. If a stimulation-evoked biomarker of antidepressant effects could be localized and monitored, it would not only improve current clinical efficacy, but even help pave the way to closed loop iterations of DBS therapy^29,36,37^.

Here, we test whether cerebro-cerebral evoked potentials (CCEPs)^38,39^ recorded from fronto-temporal regions can reveal the DBS configurations leading to a clinical improvement in patients with TRD. To this end, we retrospectively examine CCEPs obtained through single-pulse electrical stimulation (SPES) delivered through different DBS contacts in patients with TRD. Patients were part of a clinical trial with dual target SCC and VC/VS leads, in which neural activity in the frontal and temporal cortices was recorded with intracranial electrodes (by means of stereo-electroencephalography; sEEG) following DBS implantation for a 10 day period^40^ (Figure 1). Following this in-hospital monitoring period, the sEEG probes were removed and the DBS leads internalized, and the patients underwent several DBS programming sessions to identify the configurations leading to an improvement in their clinical scores (Montgomery-Åsberg Depression Rating Scale [MADRS] score^41^) before entering a double-blind phase. Here, for each patient we identified DBS configurations that led to the best clinical improvements (“best” configurations) with respect to all the tested configurations (“other” configurations), and we retrospectively analysed CCEPs obtained in response to those configurations, collected during the in-hospital monitoring period with sEEG. By assessing within-patient patterns of CCEPs associated with best and other configurations, we can determine whether a common CCEP pattern is predictive of best symptom improvement. If true, CCEPs at specific anatomical locations could be used to objectively guide device placement or aid contact selection^42^, potentially through invasive or non-invasive monitoring, such as a temporary subdural strip or scalp-EEG, eventually accelerating the achievement of a clinical improvement for patients undergoing DBS for TRD.

**Figure 1.**
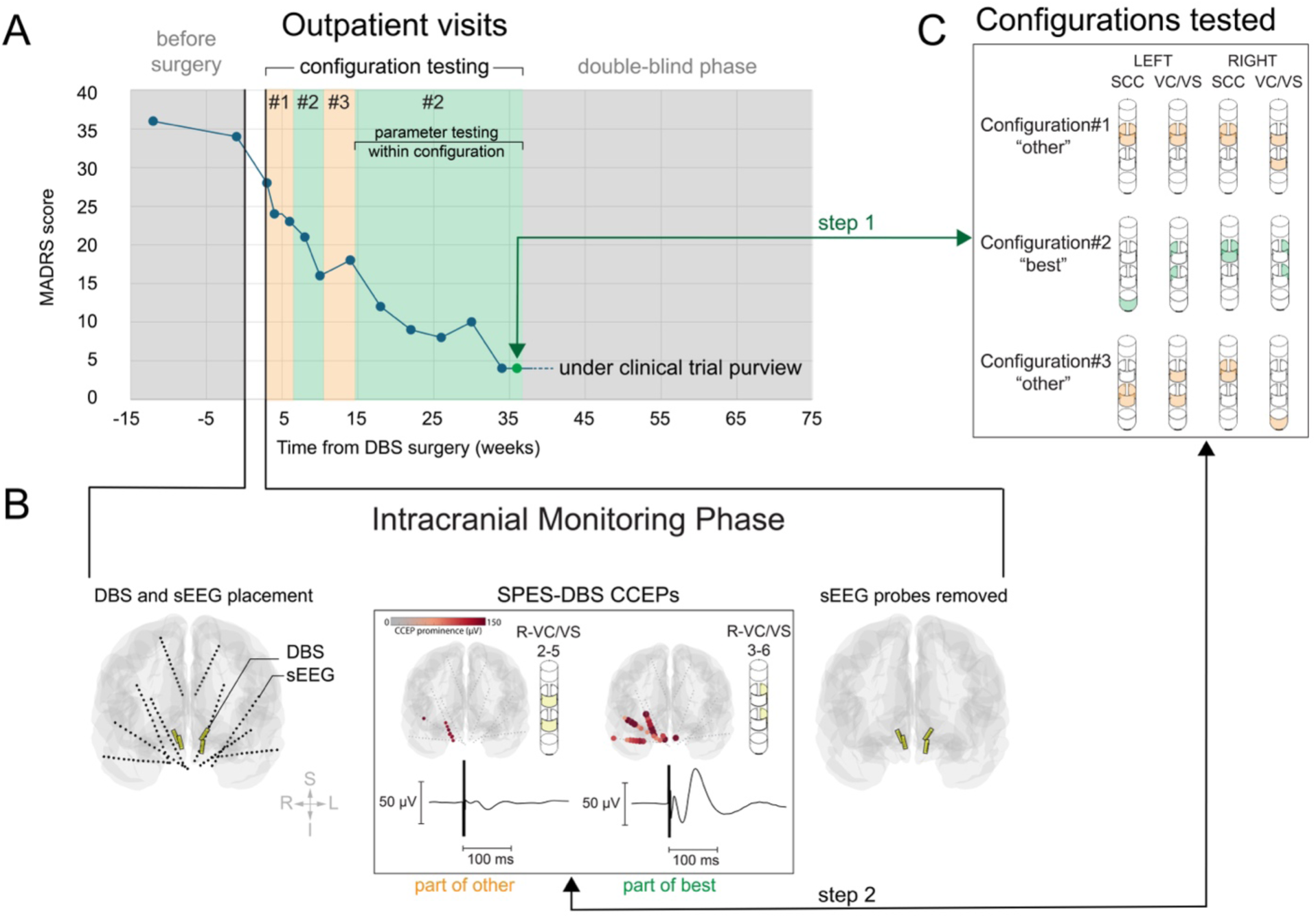
Experimental setup. Panel A: Timeline showing MADRS scores before/after DBS surgery for patient 1 and across the various phases of the trial (intracranial monitoring phase, outpatient visits with DBS configuration testing, double blind phase). The clinical results for this patient have been released and can be found in the relative publication^40^. Panel B: Zoom-in on the intracranial monitoring phase, and SPES experiments. The plots in the inset show the magnitude of the CCEPs in response to SPES delivered from DBS contacts for one patient. CCEPs are depicted at each sEEG electrode location by color-coding and inflating the size of each electrode according to the CCEP magnitude (grey to red). The average CCEP waveform is also depicted for each experiment. Panel C: Schematic of configurations tested during the outpatient visits (for this patient, three). The configuration leading to the lowest MADRS score during the outpatient visit is defined as “best” for the scope of this work. Note that the same configuration is associated with multiple MADRS scores (green areas in panel A), denoting that the same configuration was used for multiple weeks while amplitude and pulse duration parameters were varied. The arrow labelled ‘step 1’ marks the identification of the best configuration based on the lowest MADRS score (here, just before double-blind). The arrow labelled ‘step 2’ marks the retrospective identification of SPES experiments associated with stimulation of each DBS lead part of best or of other configurations (example, right VC/VS).

## Methods

### Human subjects

This study reports on data collected from eight subjects diagnosed with TRD (ages 30s-60s; four females). All are participants of an ongoing clinical trial (NCT 03437928) investigating the electrophysiological mechanisms behind the use of DBS as a therapy for TRD. All subjects provided written and informed consent for the experimental procedures part of the clinical trial, regulated by the Baylor College of Medicine IRB (H-43036). Demographic information and clinical scores for each participant will not be disclosed until the conclusion of the clinical trial (Fig. 1A).

### DBS leads and sEEG

For each subject, four DBS leads targeting bilateral SCC and VC/VS (one in each region) were surgically placed. Segmented DBS leads (Boston Scientific Cartesia) were employed for all subjects and targets, except for patients 7-8 who received 8-contact cylindrical DBS leads (Boston Scientific Vercise) for bilateral VC/VS. The Boston Scientific Cartesia DBS leads contain eight stimulation contacts: Two solid rings at the deepest and shallowest points, labelled contact 1 and 8, respectively, and two three-way segmented ring leads numbered 2-7, following the same depth convention as the solid rings. A DBS configuration is defined by which contacts of each lead are delivering stimulation (Fig. 1B). A temporary set of ten sEEG probes (PMT Corporation, MN, USA) were also placed during the same surgery, spanning fronto-temporal regions bilaterally (Fig. 1C). The sEEG probes had 0.8 mm diameter, between 8 and 16 electrode contacts along the probe length and with 3.5 mm center-to-center distance between contacts.

### Timeline and identification of best DBS configuration

After surgery, each subject was intracranially monitored for ten days at Baylor St. Luke’s Hospital (patients 1-6) or UT Southwestern Medical Center (patients 7-8). During the intracranial monitoring period (Fig. 1B), various experimental procedures were performed as part of the clinical trial. Relevant to the current work, SPES experiments were performed. After the inpatient monitoring period, patients underwent a second surgery to remove the temporary sEEG electrodes and to connect the DBS leads to implanted pulse generators (Boston Scientific Gevia). Outpatient research visits occurred on a weekly basis where configuration changes were made and clinical scores were collected. After 4 weeks, this periodicity was switched to biweekly, and then monthly (Fig. 1A-C). By visit 19 (usually occurring around 9 months after DBS implant) subjects entered a double-blind discontinuation phase. For the purpose of this work, any DBS configuration that led to the lowest MADRS score was labelled as the “best” configuration (Fig. 1C), regardless of whether the patient achieved long-term responder status (still under evaluation for some patients and not disclosed at this time, see Sheth et al., 2021 for publicly available information related to patient 1^40^).

### Electrode localization

DBS and sEEG electrode contact location was determined through the intracranial Electrode Visualization software pipeline, iELVis^43^. The postoperative clinical CT and preoperative T1 anatomical MRI were co-registered using the linear image registration tool (FLIRT) from the Functional Magnetic Resonance Imaging tool for the Brain Software Library (FMRIB^44^). Contacts were then identified on the coregistered CT in BioImage Suite^45^. The *xyz* coordinates of each electrode were used for automatic labelling based on the volumetric parcellation from the Desikan-Killiany Atlas^46^, using a radius of 6 mm around the coordinate. Anatomical labels were grouped together as follows: Orbitofrontal cortex (OFC): Lateral and medial orbitofrontal; dorsal anterior cingulate cortex (DACC): Caudal and rostral anterior cingulate, cingulate mid anterior, cingulate central; dorsal prefrontal cortex (DPFC): Caudal and rostral middle frontal, superior frontal; ventro-lateral PFC (VPFC): Pars orbitalis, triangularis and opercularis; lateral temporal cortex (LT): Middle and superior temporal cortex; medial temporal lobe (MTL): Amygdala, hippocampus and medial portions of the inferior temporal cortex; white matter (WM): Left and right white matter, with no grey matter labels detected within the 6 mm radius, to be used as a baseline for statistical analyses. Anatomical labels were visually inspected. Of note, the atlas employed does not have a dedicated label for ventromedial prefrontal cortex, and thus our OFC definition likely includes some portions of ventromedial PFC. For visualizations (Fig. 1B, Fig. 2A-B), we overlaid the electrode coordinates in MNI space on the *fsaverage* brain^47^.

**Figure 2.**
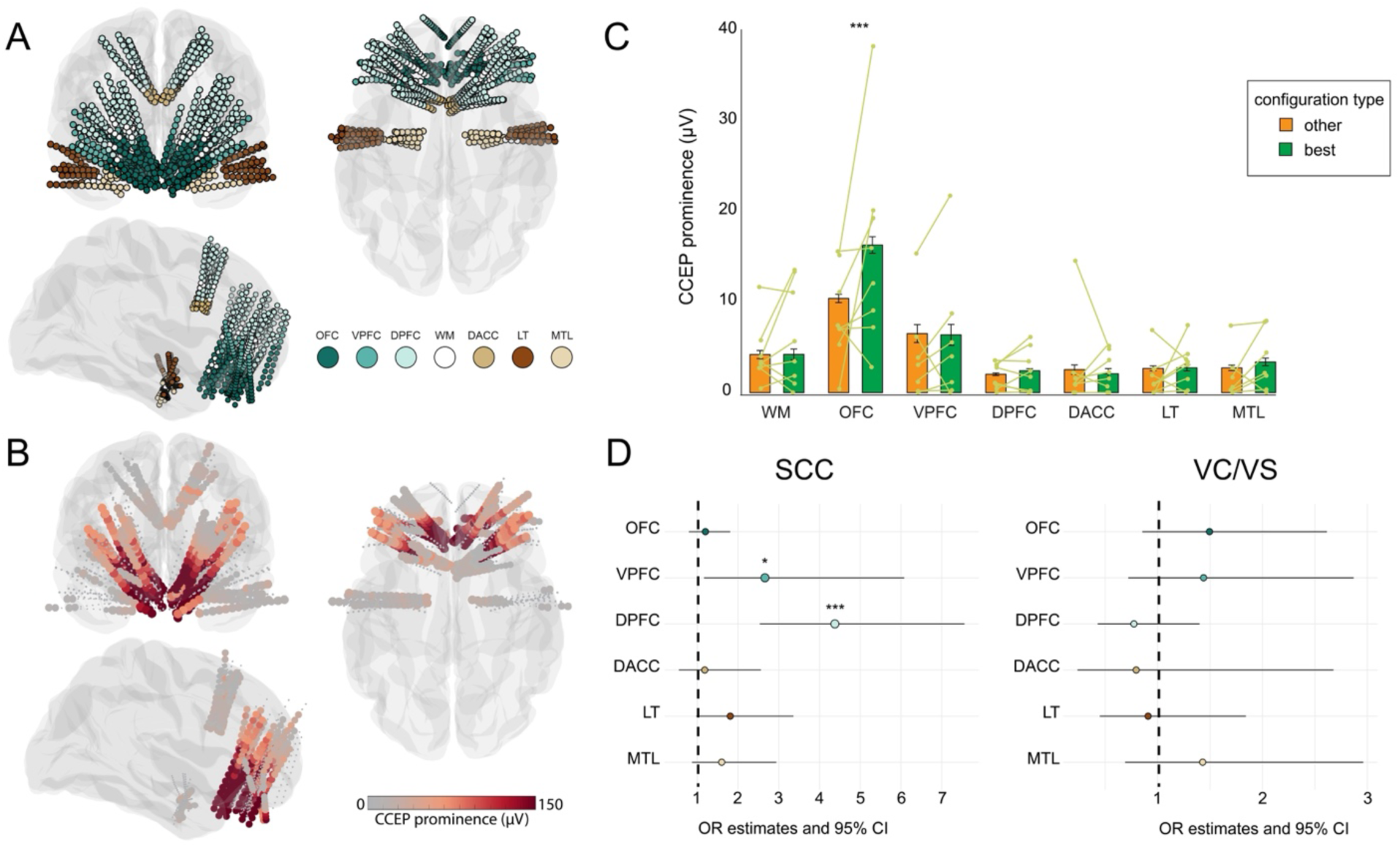
CCEPs modulations by best configuration. Panel A: Anatomical distribution of sEEG electrode contacts across all patients (MNI coordinates displayed on the *fsaverage* brain, color-coded by regions). Panel B: Electrode contacts inflated and color-coded by average CCEP peak prominence in response to stimulation of the best configurations (averaged across SCC and VC/VS best configurations). Panel C: Bar plots display mean and standard error of CCEP peak prominence for all patients divided by region and configuration type. Each line represents the patient’s average.. Panel D: Odds-ratio (OR) and confidence interval (CI) estimated from mixed-effect logistic model coefficients, for SCC and VC/VS separately, indicating change in odds of detecting a CCEP response for best versus other configurations. Asterisks denote statistical significance derived from mixed-effect models (* = *P*<0.05; *** = *P*<0.001).

### SPES-DBS stimulation parameters for CCEPs (in-patient)

Monopolar symmetric biphasic cathodic-first single pulse stimulation was delivered through the DBS leads using a Blackrock CereStim R96 stimulator (Blackrock Microsystems, Utah, USA). Stimulation consisted of 315 stimulation pulses (180 µs pulse width for each phase; 100 µs interphase gap; amplitude of 4.8 mA for all patients except patient 1, who received stimulation at 5 mA), occurring with a minimum inter-stimulation interval of 600 ms and an additional random jitter of 1-200 ms. The current return for the stimulation was a large conductive pad placed on the patient’s thigh^48^. Stimulation was delivered from one configuration at a time on a given DBS lead (i.e., each solid ring contact for each DBS lead; for segmented leads, all three contacts within the segmented rings, or all two for segmented stacks, were stimulated simultaneously with current amplitude divided accordingly). Each stimulation experiment tested one DBS configuration for a single DBS lead. Multilead configurations^49^ were not systematically tested across all patients and are not included in this work. Patients were awake for all stimulation experiments.

### Recording and preprocessing

Neural data recorded via the sEEG probes was sampled at 30 kHz using a Blackrock Microsystems NeuroPort Acquisition System (except for subject 1, sampled at 2 kHz). Ground and reference were two contacts visually confirmed to be located in white matter (patients 1-6) or two contacts from a strip electrode placed in the subgaleal space^50^ (patients 7-8). Custom pipelines were used to process the data using MATLAB (Mathworks). Electrode contacts with signal displaying artifacts and excessive line noise were excluded from all subsequent analyses (final electrode counts are reported in Table 1). Stimulation trials were epoched to 150 ms preceding and 350 ms following each stimulation pulse onset. Epoched trials were then downsampled to 2 kHz and underwent Common Average Re-referencing by Least Anticorrelation (CARLA^51,52^). All analyses were performed via custom scripts using MATLAB or R-Studio (Rstudio team, 2025).

**Table 1.** Electrode coverage information for each patient. Each row shows the number of sEEG electrode contacts in each region of interest for each patient. Only electrodes included in analysis are reported.

| Patient # | sEEG contacts across regions |  |  |  |  |  |  |  |
| --- | --- | --- | --- | --- | --- | --- | --- | --- |
|  | OFC | VPFC | DPFC | DACC | LT | MTL | WM | Total |
| 1 | 41 | 11 | 47 | 5 | 13 | 11 | 15 | 143 |
| 2 | 50 | 10 | 43 | 8 | 13 | 11 | 7 | 142 |
| 3 | 41 | 12 | 42 | 6 | 12 | - | 7 | 120 |
| 4 | 28 | 4 | 50 | 5 | 16 | 11 | 31 | 145 |
| 5 | 44 | - | 52 | 6 | 16 | 10 | 12 | 140 |
| 6 | 44 | 15 | 41 | 4 | 10 | 16 | 6 | 136 |
| 7 | 50 | 8 | 41 | 7 | 11 | 11 | 14 | 142 |
| 8 | 50 | 16 | 35 | 6 | 17 | 12 | 7 | 143 |

### MADRS scores differences for best versus other configurations

The best configuration is, by our own definition, the one leading to the lowest MADRS score. First, we wanted to confirm that the difference between best and others MADRS scores was statistically significant. We considered the whole set of outpatient visits before the double-blind phase, and we compared the MADRS scores collected during each programming visit while the patients were on the DBS configuration identified as best versus the others. Indeed, each configuration was typically used for multiple weeks and with varying stimulation settings (current amplitude, pulse width) and thus was associated with multiple MADRS scores (i.e., the best configuration was associated with multiple different scores in addition to the lowest one, Fig. 1A). We employed a mixed-effects model to test for differences between MADRS scores associated with best versus other configurations (fixed effect, with patients modelled as random effects). To further control that the difference was not solely driven by the lowest score in the best configurations, we repeated the analysis after excluding the lowest score for each patient.

### CCEP analysis

For each sEEG contact and each stimulated configuration, evoked potentials between 10-100 ms after the end of the stimulation pulse were detected using the *findpeaks* function in MATLAB on the trial-averaged trace (minimum peak prominence: 10 µV and 6 standard deviations from the pre-stimulation baseline, comparable to existing approaches^53^). Signals occurring prior to the 10 ms mark were not considered for peak detection to avoid stimulation artifact contamination. Both positive and negative deflections were considered, given that CCEPs measured with sEEG can have components at different polarities depending on the electrode position with respect to the cortical folds. If more than one peak was detected, the one with the largest absolute prominence peak value was stored for analysis. If no peak was detected, the CCEP was assigned a value of 0. The average latency and standard deviation of the detected CCEPs was 32.96±15.24 ms. The CCEP peak prominence values for all patients, contacts and configurations tested were analysed with a mixed-effects model to evaluate differences between DBS configurations (“best” versus “other” used as baseline level), while including patients as random effects. We further tested for spatial specificity by including a term identifying the anatomical locations of the contacts (OFC, VPFC, DPFC, DACC, LT, MTL and WM used as baseline level) and evaluating the interaction between each anatomical location and configuration type. Finally, we tested if the effects were dependent on the DBS lead position (SCC and VC/VS).

Next, we binarized the CCEPs as present/absent to remove the influence of their magnitude and analysed the binarized data with a mixed-effects logistic model, to detect if changes in the odds of detecting a CCEP were related to configurations type and their interaction with anatomy (patients as random effects). This was motivated by the fact that CCEPs in some anatomical regions might be smaller in magnitude, potentially hiding the presence of smaller modulations according to configuration type. Exponentiated model coefficients were used as odds-ratio (OR) estimates. Full model structure and outputs are reported in supplementary material. In the results, exact *P*-values are reported for values larger than *P*=0.001.

### PCA and similarity

In order to test for the presence of a shared spatio-temporal pattern of evoked potentials across the patient population, we performed principal component analysis (PCA) using the *pca* function in MATLAB. PCA was applied to the trial-averaged CCEP waveforms (time-series) across all recorded contacts, tested DBS configurations and patients. The first five components explained 93% of the variance. CCEP patterns obtained in response to each configuration for each DBS lead were compared to those obtained in response to the best configuration via squared euclidean distance of the scores for the first five PC using the *pdist2* function. Once calculated, the distance values of the evoked responses in PC space for each of the other configurations with respect to the best were averaged across all recorded electrode locations for each patient into a single scalar value, and then distance values were normalized within-patient between 0 (representing the best configuration) and 1 (maximum distance between the best and other configuration pairs).

To evaluate if the dissimilarity of CCEPs patterns showed an association with the clinical scores for each DBS configuration, we fitted a regression model. First, to account for differences between absolute MADRS scores across patients and for the influence of time, we computed a normalized MADRS difference score for each DBS configuration, bounded between 0 and 1 (with 0 representing the lowest MADRS score, i.e. the best) weighted by the testing duration for each DBS configuration.

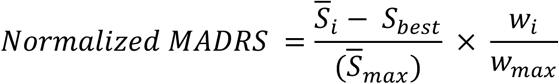

Specifically, configurations associated with more than one MADRS score first had their scores averaged (represented by *S̅_i_*). Each averaged score then had the participant’s best MADRS score subtracted from it, with the resulting sum divided by their largest averaged MADRS score (*S̅_max_*, thus placing the best and worst MADRS scores for each participant at 0 or 1, respectively). This value was then multiplied by the sum of the total weeks the configuration had been used (represented by *wi*) divided by the total weeks of the longest tested configuration (*w_max_*). This normalization mitigates the duration differences between configurations, giving less weight to configurations that were tested for shorter durations. Finally, we regressed the MADRS normalized difference against the average distance between the CCEP pattern obtained with the best configuration and every other configuration for each patient. We evaluated the slope coefficient to test for the presence of a gradient, and we performed permutation testing (5,000) and repeated the regression leaving one patient out at a time to verify statistical robustness.

### Bridging the SPES-DBS configurations from CCEPs to therapeutic outpatient settings

The stimulation parameters tested during the outpatient visits are not directly comparable with those used for the SPES experiments, thus the focus of the study is on the DBS contact configurations leading to the best MADRS score, regardless of the exact stimulation parameters employed. Indeed, the outpatient stimulation parameters use continuous high-frequency stimulation (130 Hz), and varying levels of pulse width and current amplitude values (varied during outpatient programming, range 60-180 µs and 0-9 mA) and multiple leads stimulated at the same time. Our approach relies only on the spatial configuration (i.e., the stimulated contact for each DBS lead) to match the clinical data to the CCEP data.

In one patient (patient 8) we performed an additional experiment to bridge the effects obtained in response to SPES with those associated with high-frequency stimulation. We delivered bursts of 130 Hz stimulation using trains of biphasic, symmetric pulses (same current amplitude and pulse duration parameters as the ones used for SPES; 20 pulses in each train, lasting 154 ms, repeated 90 times and spaced 1.3-1.7 seconds apart). We quantified the average evoked potentials after the bursts using the root mean square (RMS) of the potential values over the 10-100 ms time window after the stimulation. We evaluated its spatial similarity to the evoked potentials in response to SPES (quantified in the same way) using correlations across all electrode locations.

## Results

### Best DBS configurations are associated with lower MADRS scores

When considering the whole set of MADRS scores collected during outpatient visits (Fig. 1A), we confirmed that best configurations resulted in MADRS scores that were significantly lower than other tested configurations (β = 8.34, *P*<0.001). This was true also when excluding the lowest score for each patient (β = 6.28, *P*<0.001), confirming that patients had overall better symptom improvement while on the best configuration regardless of specific DBS parameters used and also without considering the score used to identify the configuration as best.

### Best DBS configurations yield stronger and more prevalent CCEPs in OFC

Stimulation delivered from DBS contacts which were later identified as part of the best configuration evoked significantly larger CCEPs with respect to stimulation from other configurations (β =2.46, *P*<0.001). When visualized, large bilateral frontal CCEPs were evident (Fig. 2B). We next assessed the spatial specificity of this effect by introducing the anatomical location term and its interaction with configuration type (best vs other), which revealed the effect to be located in the OFC (mean and standard error of the mean for OFC CCEPs associated with best configuration = 16.3±0.92 µV, with other configuration = 10.4±0.47 µV; interaction between configuration type and CCEPs in OFC: β = 6.54, *P*<0.001; Fig. 2C). No other anatomical location displayed a reliable modulation of CCEPs according to the configuration type (i.e., no other interactions). The effect was not dependent on any specific patient, assessed by re-running the model while leaving one patient out at a time (interaction between configuration type and OFC CCEPs ranging between 4.8 and 8.01, all *P*<0.001; no other significant interaction terms). The interaction with best configuration was present for both medial and lateral OFC subdivisions (medial: β = 6.98, P<0.001; lateral: β = 6.21, P<0.001; supplementary material).

Finally, we introduced a model term to separate each DBS lead location (SCC and VC/VS), and this revealed that the DBS lead was modulating the increase in OFC CCEPs magnitude for the best configuration (three-way interaction between configuration type, CCEPs in OFC and DBS lead: β = 6.88, p=0.003). To tease apart the interaction, we re-ran the models for SCC and VC/VS separately, and we confirmed that for both DBS locations the interaction between best configurations and OFC CCEPs was significant, although larger for SCC (interaction between configuration type and CCEPs in OFC in SCC: β = 8.4, *P<*0.001; in VC/VS: β = 2.52, p=0.024). No anatomical locations other than OFC were associated with CCEP modulations by best configurations for either SCC or VC/VS.

### Higher odds of prefrontal CCEPs for best SCC configurations

Next, we analysed the binarized data to remove the influence of CCEP magnitude (CCEP present/absent). The change in the odds of detecting a CCEP for best versus other configurations was significant only in OFC (β = 0.38, p=0.023, OR 1.46) and no other location (all p>0.05), resembling the effect obtained when analysing CCEP magnitude. However, when we ran the model separately for SCC and VC/VS, we identified specific patterns that did not reach statistical significance when considering them together. The best SCC configurations were associated with increased odds of CCEPs in DPFC (β = 1.48, *P*<0.001, OR 4.38) and VPFC (β = 0.98, p=0.02, OR 2.66). On the contrary, VC/VS configurations did not show any specific changes in the odds of detecting CCEPs for best versus other configurations at any location (no significant interaction terms, all p>0.05). Thus, when removing the influence of the CCEP magnitude and focusing only on their presence/absence, dorsal and ventro-lateral PFC locations displayed increased numbers of CCEPs for best SCC configurations (Fig. 2D).

### Even non-optimal DBS configurations lead to symptom improvement if their CCEPs are similar to those elicited by the best configurations

To test whether CCEP patterns could explain the spectrum of clinical responses from the other configurations (not best), we computed the distance between all other CCEP patterns and the best one in PC space for each patient (Fig. 3A) and regressed that distance against MADRS score differences. The group-level data displayed a positive relationship between distance from the best CCEP pattern and their MADRS scores (R^2^ = 0.43; intercept = 0.041, p = 0.4; slope = 0.294, *P*<0.001), indicating that CCEP patterns closer to the best were associated with better MADRS scores, even for non-optimal configurations (Fig. 3B). We further validated the result of a spectrum between CCEP patterns and clinical improvement with permutation testing by shuffling the association between MADRS scores and CCEP distance values (null hypothesis: Slope not different from 0; p = 0.008) and by re-running the regression model with a leave one out approach (removing a patient’s configuration one at a time to ensure the effect was not driven by any given patient/configuration: Slope min = 0.267; max = 0.337, all *P*<0.001).

**Figure 3.**
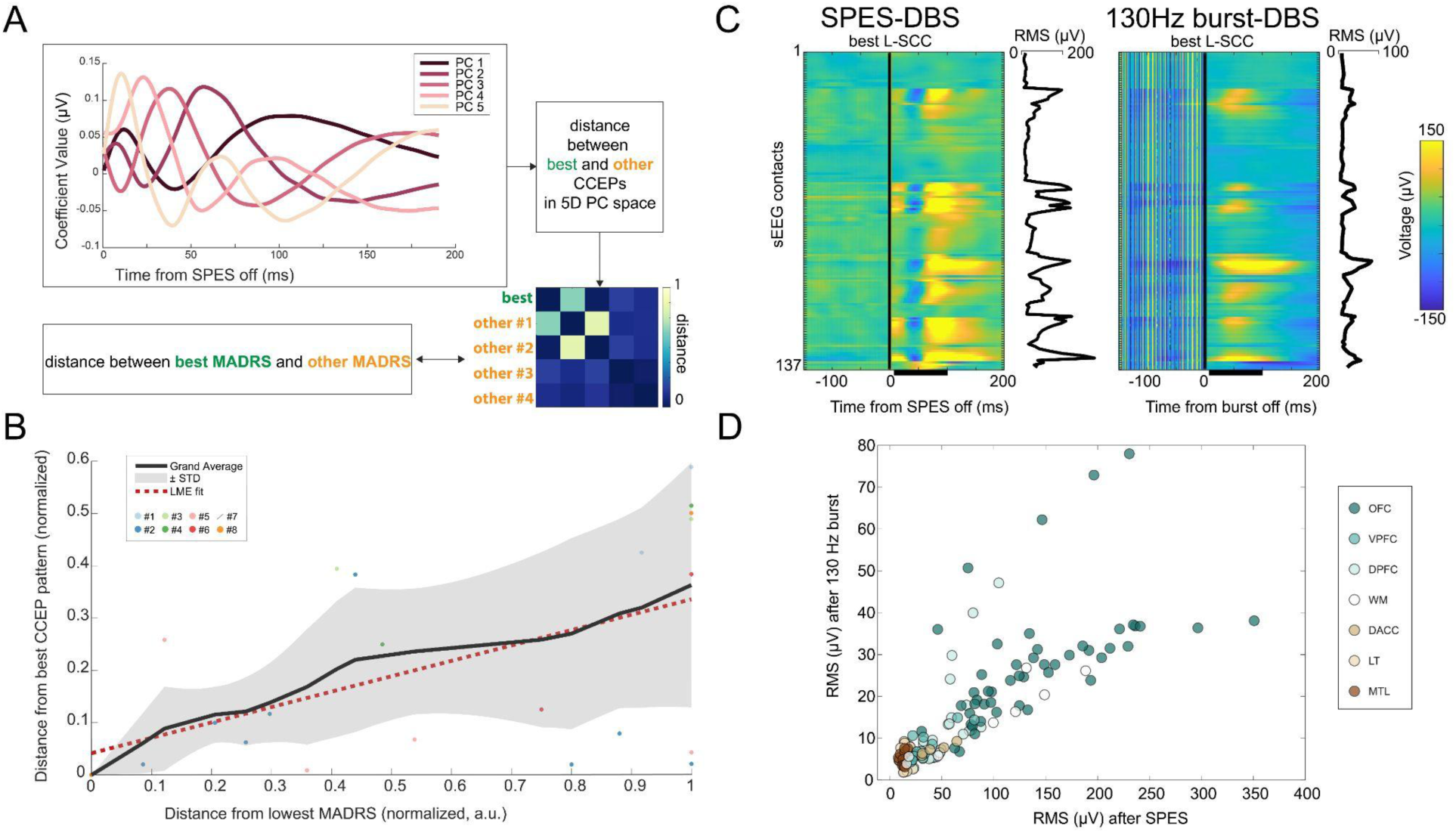
Relationship between CCEP patterns and therapeutic effects. Panel A: Schematic showing the computation of similarity between the CCEP patterns evoked by various configurations. On top, the first five principal components obtained across all CCEP data. Distance between CCEPS from best and other configurations is computed in the five dimensional space for each patient and each SPES-DBS. On the right, the average distance values for patient 6 are displayed as an example. The distance values are then compared to the distance between best and other MADRS scores. Panel B: Correlation between CCEP patterns and MADRS scores (distances). CCEP patterns more closely resembling those obtained in response to the best configuration are associated with better MADRS scores. Black line is the grand average across patients and configurations, and the grey area shows the standard deviation. The regression fit is shown overlaid as a red-dashed line, each point represents a patient’s tested configuration. Panel C: CCEPs after SPES-DBS (left) and after 130 Hz bursts (right), showing changes in voltage across all sEEG electrodes (averaged across repeats of stimulation). The RMS values computed from 10-100 ms after the stimulation are displayed on the right side of each plot. Panel D: CCEPs RMS values for SPES and 130 Hz burst, color-coded by anatomical region, showcasing the similarity between the two sets of values, with OFC and DPFC locations displaying the strongest responses to the high-frequency bursts.

### Similar spatial distribution of evoked responses to single pulse and high frequency stimulation

We compared the CCEPs evoked by SPES and by bursts of 130 Hz stimulation in a single patient (patient 8). We found that CCEPs evoked from short-bursts of 130 Hz stimulation were spatially similar to those evoked by SPES (r = 0.78, *P*<0.001) delivered through the same DBS contact, indicating that the effect of high-frequency stimulation on interconnected regions might be tested using short bursts of stimulation or using SPES (Fig. 3C). While the spatial pattern of the evoked responses was similar, their magnitude was lower for the bursts. Interestingly, a few OFC and DPFC locations displayed higher-than-expected RMS values in response to the bursts, bifurcating from the main pattern (Fig. 3D).

## Discussion

Determining why DBS therapy outcomes for neuropsychiatric disorders remain variable, as well as why certain DBS configurations outperform others, continue to be large hurdles for the standard implementation of this treatment for refractory conditions^54,55^. The temporal delay between the start of DBS treatment and measurable clinical changes in depressive symptoms is a major roadblock to the advancement of therapeutic options for patients with TRD, as the parameter space is vast and cannot be empirically tested for each patient. The present study contributes to the growing body of work attempting to overcome these obstacles^56–60^. By combining long-term clinical assessments with retrospective examination of acute, stimulation-driven biomarkers, we demonstrate that the most effective SCC and VC/VS DBS configurations were associated with large CCEPs in the OFC and with higher odds of CCEPs in dorsal and ventro-lateral prefrontal cortex, offering a potential stimulation-driven biomarker to guide contact selection for DBS in TRD patients. The therapeutic effects of DBS were not binary but rather graded, suggesting that there is a configuration subspace with multiple potential DBS configurations capable of obtaining similar symptom improvement.

The association between OFC response and DBS effectiveness is supported by the central role of OFC in the pathophysiology of depression, with converging evidence showing that both its local activity and connectivity profile contribute to core symptoms. OFC dysfunction is associated with anhedonia, rumination, blunted reward sensitivity, increased loss anticipation and heightened negative bias^61–71^. In addition to OFC engagement, we found that the most effective SCC DBS configurations were also associated with increased odds of CCEPs in the DPFC, and to a smaller extent in the lateral VPFC. Dorsal and ventrolateral PFC are implicated in cognitive control dysfunctions in depression, contributing to emotional dysregulation and difficulties in decision making^72,73^.

Effective neuromodulation, as currently understood, has the ability to stabilize or mitigate the effects of aberrant neural circuits through anatomical and functional connections^11,74–76^. Both the SCC and VC/VS targets are located at the confluence of axons from many different PFC regions. The precise electrode location, within each patient’s individual anatomy, determines which axons are impacted by stimulation. Ipsilaterally, all of the pathways described here are plausibly (although not necessarily) monosynaptic in nature. The OFC, VPFC and DPFC all send fibers through the VC to reach the thalamus, subthalamic nucleus and brainstem^77,78^; the OFC and VPFC also send terminating axons to the nearby VS^79^. The OFC also projects directly to the grey matter of the SCC and through the white matter of the subgenual cingulum bundle adjacent to it, while DPFC and VPFC fibers traverse the forceps minor at the SCC target^56,80^. Strong prefrontal CCEPs in response to DBS-SPES could constitute an objective way to evaluate if any given DBS contact or configuration is capable of optimally engaging the circuit of interest^53^, and thus more likely to be effective therapeutically^30^.

The distance analysis between best configurations and other configurations revealed the presence of a gradient, such that depressive symptoms improvement scaled with the similarity of CCEPs to the best configuration. This finding indicates how the ideal stimulation-evoked state achieving the best possible clinical response is not an all-or-nothing dichotomy, but this exists on a continuum, with multiple configurations capable of reaching similar and intermediate clinical improvements. Previous studies utilizing electrophysiological signals for DBS therapeutic guidance in movement disorders have observed similar phenomena^32,81^. For example, Xu et al. (2022) investigated the efficacy of using intraoperative ERNA to guide DBS contact selection for Parkinson’s disease and found similar clinical efficacy between those guided by the neural responses and those selected by physicians based on symptom improvement. Furthermore, the benefit gained by those electrophysiological contacts (most ideal, then second most ideal, then third, etc.) behaved in a graded way, similar to our results here. Electrophysiological signals able to perform as well as more traditionally accepted forms of guidance, such as imaging- and symptom-guided, while identifying and ranking multiple potential configurations, are uniquely posed to accelerate contact selection and improve the efficacy of DBS therapy^82–85^. Indeed, acute measures of objective DBS-SPES evoked activity could help determine the configuration space most likely to achieve a symptom improvement, limiting the trial-and-error approach for finding an effective configuration during the outpatient period^81^. Furthermore, chronic monitoring of symptoms fluctuations and side effects can be used to refine personalization among the available configurations, widening the therapeutic window while minimizing side effects.

One open question is whether the effects of single-pulse stimulation can be used to make inferences of the neural circuitry mediating the therapeutic response to DBS. Here, we leverage CCEPs from single-pulse stimulation to identify the signatures that distinguish DBS contact configurations associated with the best clinical improvement from the others. However, aside from their practical predictive value, it is not clear if we can assume that therapeutic DBS settings engage the circuitry in a similar manner. Indeed, clinical applications of DBS utilize a high frequency stimulation parameter (usually between 130 and 180 Hz^86^). SPES is a significant departure from clinical DBS parameters, and it is not well established if neural responses to SPES are representative of those during DBS therapy. A similar assumption has been recently used to investigate the effect of DLPFC-TMS for TRD on limbic and striato-thalamic locations^87^. SPES is used to empirically reveal regions displaying strong connectivity to the stimulated location^88^, and it is likely that long-term high frequency stimulation may achieve its therapeutic effect by modulating the activity of interconnected regions, as recently demonstrated in DBS for epilepsy^89^. However, there is limited evidence directly comparing neural responses and clinical translatability of single pulses versus high-frequency stimulation. A recent study^90^ found that the neural responses generated by STN stimulation at 10 Hz and 130 Hz were highly correlated. Similarly, our results suggest a strong correlation between the spatial patterns of activity evoked by SPES and by bursts of stimulation at the target therapeutic frequency (here, 130 Hz), indicating that both approaches may be used to verify engagement of the circuit of interest.

While providing similar information in terms of engaged circuitry, the CCEP response to the 130 Hz bursts showed some interesting differences with respect to SPES that will require future investigations to be fully characterized. CCEPS were lower in magnitude for 130 Hz bursts versus SPES, which could reflect habituation effects. Interestingly, some OFC and DPFC locations displayed a higher-than-expected CCEP for the 130 Hz bursts, indicating that there might be location-specific tuning to different stimulation frequencies^91^. Our comparison, coming from a single patient, is too limited to fully disentangle these differences, but it provides critical evidence of the need for a deeper, mechanistic understanding of the effects of different stimulation parameters on local and interconnected brain regions.

Future studies aiming at advancing personalized neuromodulation approaches for TRD should include systematic evaluation of acute intraoperative biomarkers. Based on our results, acute monitoring of OFC and lateral PFC activity should be prioritized. This could be achieved through temporary subdural strips placed though the cranial opening used for the DBS implant^92,93^, or through non-invasive scalp-EEG monitoring combined with source analysis^35,57,85^. Importantly, future studies should investigate the precise source of the OFC biomarker, as the operational definition of OFC in the present work spans both medial and lateral portions, as well as some portions of ventromedial PFC. Although we did not find medial-lateral differences (Supplementary analysis), dedicated studies would be needed to disentangle the contribution of each anatomical subdivision. Here, we leveraged a retrospective approach to identify CCEPs associated with configurations leading to better symptom improvement, but prospective, systematic evaluations with larger samples will be necessary to define objective guidelines to incorporate SPES-biomarkers in DBS surgical practice. One key aspect of our study is the reliance on within-patient comparisons for the majority of the analyses. Our approach is based on within-patient differences, specifically between configurations leading to different levels of symptom improvement, and their consistency at the group level. This approach avoids confounds arising from inter-individual differences, which affects the common approach of comparing effects between subgroups of responders versus non-responders, especially critical when dealing with the small sample sizes typical of neuropsychiatric DBS cohorts.

Existing data-driven neuromodulation approaches already recognize the importance of using individualized anatomical connectivity to improve treatment efficacy^94^. The development of functional, symptom-based^95,96^ or stimulation-based personalized approaches^97^ may identify new therapeutic options for treatment resistant conditions^98^. Indeed, stimulation-based electrophysiological features indicative of circuit-engagement are not only posed to reduce clinical efficacy variability, but they will also provide mechanistic information on the fundamental pathways mediating the therapeutic response^99^. Thus, identifying objective, acute and anatomically-specific biomarkers is essential to reduce therapeutic variability and accelerate clinical efficacy for the next-generation of data-driven personalized neuromodulation approaches^98^.

Altogether, our work supports the use of prefrontal CCEPs to guide contact selection in DBS for TRD and underscores the importance of further, dedicated investigation into OFC acute biomarkers.

## Data availability

The data for this study is not publicly available to protect the privacy of participants who are a part of an ongoing clinical trial (NCT 03437928). Contact the corresponding author for reasonable requests and inquiries about this dataset.

## References

1. Key Substance Use and Mental Health Indicators in the United States: Results from the 2021 National Survey on Drug Use and Health.

2. Bains N, Abdijadid S. Major Depressive Disorder. In: StatPearls. StatPearls Publishing; 2025. Accessed March 4, 2026. http://www.ncbi.nlm.nih.gov/books/NBK559078/

3. Malhi GS, Mann JJ. Depression. The Lancet. 2018;392(10161):2299-2312. doi:10.1016/S0140-6736(18)31948-2

4. Sun J, Ma Y, Guo C, et al. Distinct patterns of functional brain network integration between treatment-resistant depression and non treatment-resistant depression: A resting-state functional magnetic resonance imaging study. Prog Neuropsychopharmacol Biol Psychiatry. 2023;120:110621. doi:10.1016/j.pnpbp.2022.110621

5. Javaheripour N, Li M, Chand T, et al. Altered resting-state functional connectome in major depressive disorder: a mega-analysis from the PsyMRI consortium. Transl Psychiatry. 2021;11(1):511. doi:10.1038/s41398-021-01619-w

6. Rush AJ, Trivedi MH, Wisniewski, SR, et al. Acute and Longer-Term Outcomes in Depressed Outpatients Requiring One or Several Treatment Steps: A STAR*D Report. Am J Psychiatry. Accessed March 4, 2026. https://psychiatryonline.org/doi/10.1176/ajp.2006.163.11.1905

7. Hasin DS, Sarvet AL, Meyers JL, et al. Epidemiology of Adult DSM-5 Major Depressive Disorder and Its Specifiers in the United States. JAMA Psychiatry. 2018;75(4):336–346. doi:10.1001/jamapsychiatry.2017.4602

8. McIntyre RS, Alsuwaidan M, Baune BT, et al. Treatment-resistant depression: definition, prevalence, detection, management, and investigational interventions. World Psychiatry. 2023;22(3):394–412. doi:10.1002/wps.21120

9. Lozano AM, Giacobbe P, Hamani C, et al. A multicenter pilot study of subcallosal cingulate area deep brain stimulation for treatment-resistant depression. J Neurosurg. 2012;116(2):315–322. doi:10.3171/2011.10.JNS102122

10. Riva-Posse P, Choi SK, Holtzheimer PE, et al. A connectomic approach for subcallosal cingulate deep brain stimulation surgery: prospective targeting in treatment-resistant depression. Mol Psychiatry. 2018;23(4):843–849. doi:10.1038/mp.2017.59

11. Reddy S, Kabotyanski KE, Hirani S, et al. Efficacy of Deep Brain Stimulation for Treatment-Resistant Depression: Systematic Review and Meta-Analysis. Biol Psychiatry Cogn Neurosci Neuroimaging. 2024;9(12):1239–1248. doi:10.1016/j.bpsc.2024.08.013

12. Gouveia FV, Warsi NM, Suresh H, Matin R, Ibrahim GM. Neurostimulation treatments for epilepsy: Deep brain stimulation, responsive neurostimulation and vagus nerve stimulation. Neurotherapeutics. 2024;21(3):e00308. doi:10.1016/j.neurot.2023.e00308

13. Fox SH, Katzenschlager R, Lim SY, et al. International Parkinson and movement disorder society evidence-based medicine review: Update on treatments for the motor symptoms of Parkinson’s disease - Fox - 2018 - Movement Disorders - Wiley Online Library. Accessed March 5, 2026. https://movementdisorders.onlinelibrary.wiley.com/doi/full/10.1002/mds.27372?saml_referrer

14. Hopfner F, Deuschl G. Managing Essential Tremor. Neurotherapeutics. 2020;17(4):1603-1621. doi:10.1007/s13311-020-00899-2

15. Dastin-van Rijn EM, Mensinger M, Dougherty DD, Widge AS. Chapter 9 - DBS in psychiatric disorders. In: Jahanshahi A, Benazzouz A, Temel Y, eds. The Neuroscience of Deep Brain Stimulation. Academic Press; 2026:209–270. doi:10.1016/B978-0-323-99742-3.00015-1

16. Johnson KA, Okun MS, Scangos KW, Mayberg HS, de Hemptinne C. Deep brain stimulation for refractory major depressive disorder: a comprehensive review. Mol Psychiatry. 2024;29(4):1075–1087. doi:10.1038/s41380-023-02394-4

17. Mayberg HS, Lozano AM, Voon V, et al. Deep Brain Stimulation for Treatment-Resistant Depression. Neuron. 2005;45(5):651–660. doi:10.1016/j.neuron.2005.02.014

18. Lozano AM, Mayberg HS, Giacobbe P, Hamani C, Craddock RC, Kennedy SH. Subcallosal Cingulate Gyrus Deep Brain Stimulation for Treatment-Resistant Depression. Biol Psychiatry. 2008;64(6):461–467. doi:10.1016/j.biopsych.2008.05.034

19. Brown EC, Clark DL, Forkert ND, Molnar CP, Kiss ZHT, Ramasubbu R. Metabolic activity in subcallosal cingulate predicts response to deep brain stimulation for depression. Neuropsychopharmacology. 2020;45(10):1681–1688. doi:10.1038/s41386-020-0745-5

20. Conen S, Matthews JC, Patel NK, Anton-Rodriguez J, Talbot PS. Acute and chronic changes in brain activity with deep brain stimulation for refractory depression. J Psychopharmacol (Oxf). 2018;32(4):430–440. doi:10.1177/0269881117742668

21. Adkinson JA, Tsolaki E, Sheth SA, et al. Imaging versus electrographic connectivity in human mood-related fronto-temporal networks. Brain Stimulat. 2022;15(3):554–565. doi:10.1016/j.brs.2022.03.002

22. Rauch SL, Dougherty DD, Malone D, et al. A functional neuroimaging investigation of deep brain stimulation in patients with obsessive–compulsive disorder. J Neurosurg. 2006;104(4):558–565. doi:10.3171/jns.2006.104.4.558

23. Bourne SK, Eckhardt CA, Sheth SA, Eskandar EN. Mechanisms of deep brain stimulation for obsessive compulsive disorder: effects upon cells and circuits. Front Integr Neurosci. 2012;6:29. doi:10.3389/fnint.2012.00029

24. Elias GJB, Germann J, Boutet A, et al. Local neuroanatomical and tract-based proxies of optimal subcallosal cingulate deep brain stimulation. Brain Stimul Basic Transl Clin Res Neuromodulation. 2023;16(5):1259–1272. doi:10.1016/j.brs.2023.08.014

25. Sadeghzadeh S, Swaminathan A, Bhanot P, et al. Emerging Outlook on Personalized Neuromodulation for Depression: Insights From Tractography-Based Targeting. BiolPsychiatry Cogn Neurosci Neuroimaging. 2024;9(8):754–764. doi:10.1016/j.bpsc.2024.04.007

26. Cattarinussi G, Moghaddam HS, Aarabi MH, et al. White Matter Microstructure Associated with the Antidepressant Effects of Deep Brain Stimulation in Treatment-Resistant Depression: A Review of Diffusion Tensor Imaging Studies. Int J Mol Sci. 2022;23(23):15379. doi:10.3390/ijms232315379

27. Li N, Baldermann JC, Kibleur A, et al. A unified connectomic target for deep brain stimulation in obsessive-compulsive disorder. Nat Commun. 2020;11:3364. doi:10.1038/s41467-020-16734-3

28. Holt AB, Kormann E, Gulberti A, et al. Phase-Dependent Suppression of Beta Oscillations in Parkinson’s Disease Patients. J Neurosci. 2019;39(6):1119–1134. doi:10.1523/JNEUROSCI.1913-18.2018

29. Wiest C, Simpson TG, Pogosyan A, et al. Stimulation-Evoked Resonant Neural Activity in the Subthalamic Nucleus Is Modulated by Sleep. Mov Disord. 2025;40(2):351–356. doi:10.1002/mds.30063

30. Tsai K, Cole ER, Nitcheu GLT, et al. Applications of Single-Pulse Evoked Potential Mapping for Therapeutic Brain Stimulation: A Systematic Review. Medicine and Pharmacology. Preprint posted online May 27, 2026. doi:10.20944/preprints202605.1875.v1

31. Steiner LA, Crompton D, Sumarac S, et al. Neural signatures of indirect pathway activity during subthalamic stimulation in Parkinson’s disease. Nat Commun. 2024;15(1):3130. doi:10.1038/s41467-024-47552-6

32. Xu SS, Sinclair NC, Bulluss KJ, et al. Towards guided and automated programming of subthalamic area stimulation in Parkinson’s disease. Brain Commun. 2022;4(1):fcac003. doi:10.1093/braincomms/fcac003

33. Wiest C, He S, Duchet B, et al. Evoked Resonant Neural Activity in subthalamic local field potentials reflects basal ganglia network dynamics. Neurobiol Dis. 2023;178:106019. doi:10.1016/j.nbd.2023.106019

34. Fitoz EC, Alagapan S, Cha J, et al. Common Electrophysiology Biomarkers Collected at Home Robustly Track Depression Recovery With Deep Brain Stimulation. medRxiv. Preprint posted online April 20, 2026:2026.04.13.26350107. doi:10.64898/2026.04.13.26350107

35. Waters AC, Veerakumar A, Choi KS, et al. Test–retest reliability of a stimulation-locked evoked response to deep brain stimulation in subcallosal cingulate for treatment resistant depression. Hum Brain Mapp. 2018;39(12):4844–4856. doi:10.1002/hbm.24327

36. Ansó J, Benjaber M, Parks B, et al. Concurrent stimulation and sensing in bi-directional brain interfaces: a multi-site translational experience. J Neural Eng. 2022;19(2):10.1088/1741-2552/ac59a3. doi:10.1088/1741-2552/ac59a3

37. Little S, Brown P. What brain signals are suitable for feedback control of deep brain stimulation in Parkinson’s disease? Ann N Y Acad Sci. 2012;1265(1):9–24. doi:10.1111/j.1749-6632.2012.06650.x

38. Lyu D, Stiger JR, Lusk Z, Buch V, Parvizi J. Mapping human thalamocortical connectivity with electrical stimulation and recording. Nat Neurosci. 2025;28(8):1797–1809. doi:10.1038/s41593-025-02009-x

39. Keller CJ, Honey CJ, Mégevand P, Entz L, Ulbert I, Mehta AD. Mapping human brain networks with cortico-cortical evoked potentials. Philos Trans R Soc B Biol Sci. 2014;369(1653):20130528. doi:10.1098/rstb.2013.0528

40. Sheth SA, Bijanki KR, Metzger B, et al. Deep brain stimulation for depression informed by intracranial recordings. Biol Psychiatry. 2022;92(3):246–251. doi:10.1016/j.biopsych.2021.11.007

41. Montgomery SA, Åsberg M. A New Depression Scale Designed to be Sensitive to Change. Br J Psychiatry. 1979;134(4):382–389. doi:10.1192/bjp.134.4.382

42. Horn A, Neumann WJ. From adaptive deep brain stimulation to adaptive circuit targeting. Nat Rev Neurol. 2025;21(10):556–566. doi:10.1038/s41582-025-01131-5

43. Groppe DM, Bickel S, Dykstra AR, et al. iELVis: An open source MATLAB toolbox for localizing and visualizing human intracranial electrode data. J Neurosci Methods. 2017;281:40–48. doi:10.1016/j.jneumeth.2017.01.022

44. Jenkinson M, Smith S. A global optimisation method for robust affine registration of brain images. Med Image Anal. 2001;5(2):143–156. doi:10.1016/S1361-8415(01)00036-6

45. Papademetris X, Jackowski MP, Rajeevan N, et al. BioImage Suite: An integrated medical image analysis suite: An update. Insight J. 2006;2006:209.

46. Desikan RS, Ségonne F, Fischl B, et al. An automated labeling system for subdividing the human cerebral cortex on MRI scans into gyral based regions of interest. NeuroImage. 2006;31(3):968–980. doi:10.1016/j.neuroimage.2006.01.021

47. Fischl B. FreeSurfer. NeuroImage. 2012;62(2):774-781. doi:10.1016/j.neuroimage.2012.01.021

48. Oswalt D, Beauchamp MS, Isaac Chen HC, Yoshor D. Reducing artifacts during human intracranial electrical stimulation by separating current return from recording ground. Brain Stimulat. 2025;18(6):2040–2042. doi:10.1016/j.brs.2025.10.017

49. Ghazavi A, Mirpour K, Soroush M, et al. Combinatorial effects of multi-site stimulation on depression-related brain regions: clinical data analysis and predictive modeling. Front Psychiatry. 2026;17. doi:10.3389/fpsyt.2026.1808486

50. Devara D, Jourahmad Z, Paulo DL, et al. Subgaleal versus white matter reference for stereo-electroencephalography recordings: comparison of clinical impact and signal quality. J Neurosurg. 2026;1(aop):1–11. doi:10.3171/2025.12.JNS251675

51. Huang H, Valencia GO, Gregg NM, et al. CARLA: Adjusted common average referencing for cortico-cortical evoked potential data. J Neurosci Methods. 2024;407:110153. doi:10.1016/j.jneumeth.2024.110153

52. Huang H, Adkinson JA, Jensen MA, et al. Proper reference selection and re-referencing to mitigate bias in single pulse electrical stimulation data. bioRxiv. Published online October 22, 2024:2024.10.21.619449. doi:10.1101/2024.10.21.619449

53. Avalos-Alais S, Jedynak M, Boyer A, et al. High-resolution electrophysiological mapping of effective connectivity of lateral prefrontal cortex. Brain. 2026;149(3):963–975. doi:10.1093/brain/awaf317

54. Soto Joven JT, Bermudez Riveros CC, Alzate Granados JP. Deep brain stimulation in neuropsychiatric disorders: A systematic review. Neurol Perspect. 2026;6(1):100209. doi:10.1016/j.neurop.2025.100209

55. Lozano AM, Lipsman N, Bergman H, et al. Deep brain stimulation: current challenges and future directions. Nat Rev Neurol. 2019;15(3):148–160. doi:10.1038/s41582-018-0128-2

56. Riva-Posse P, Choi KS, Holtzheimer PE, et al. Defining Critical White Matter Pathways Mediating Successful Subcallosal Cingulate Deep Brain Stimulation for Treatment-Resistant Depression. Biol Psychiatry. 2014;76(12):963–969. doi:10.1016/j.biopsych.2014.03.029

57. Seas A, Noor MS, Choi KS, et al. Subcallosal cingulate deep brain stimulation evokes two distinct cortical responses via differential white matter activation. Proc Natl Acad Sci U S A. 121(14):e2314918121. doi:10.1073/pnas.2314918121

58. Sheth SA, Mayberg HS. Deep Brain Stimulation for Obsessive-Compulsive Disorder and Depression. Annu Rev Neurosci. 2023;46(1):341–358. doi:10.1146/annurev-neuro-110122-110434

59. Scangos KW, Khambhati AN, Daly PM, et al. Closed-loop neuromodulation in an individual with treatment-resistant depression. Nat Med. 2021;27(10):1696–1700. doi:10.1038/s41591-021-01480-w

60. Starkweather CK, Sugrue LP, Cajigas I, et al. Stereoelectroencephalography Electrode Implantation for Inpatient Workup of Treatment-Resistant Depression. Neurosurgery. 2024;95(4):941–948. doi:10.1227/neu.0000000000002942

61. Drevets WC. Neuroimaging studies of mood disorders. Biol Psychiatry. 2000;48(8):813–829. doi:10.1016/S0006-3223(00)01020-9

62. Drevets WC. Orbitofrontal Cortex Function and Structure in Depression. Ann N Y Acad Sci. 2007;1121(1):499-527. doi:10.1196/annals.1401.029

63. Zhang B, Rolls ET, Wang X, Xie C, Cheng W, Feng J. Roles of the medial and lateral orbitofrontal cortex in major depression and its treatment. Mol Psychiatry. 2024;29(4):914–928. doi:10.1038/s41380-023-02380-w

64. Fan X, Mocchi M, Pascuzzi B, et al. Brain mechanisms underlying the emotion processing bias in treatment-resistant depression. Nat Ment Health. 2024;2(5):583–592. doi:10.1038/s44220-024-00238-w

65. Willinger D, Karipidis II, Neuer S, et al. Maladaptive Avoidance Learning in the Orbitofrontal Cortex in Adolescents With Major Depression. Biol Psychiatry Cogn Neurosci Neuroimaging. 2022;7(3):293–301. doi:10.1016/j.bpsc.2021.06.005

66. Pizzagalli DA, Roberts AC. Prefrontal cortex and depression. Neuropsychopharmacology. 2022;47(1):225-246. doi:10.1038/s41386-021-01101-7

67. Calzà J, Gürsel DA, Schmitz-Koep B, et al. Altered Cortico–Striatal Functional Connectivity During Resting State in Obsessive–Compulsive Disorder. Front Psychiatry. 2019;10:319. doi:10.3389/fpsyt.2019.00319

68. Peters SK, Dunlop K, Downar J. Cortico-Striatal-Thalamic Loop Circuits of the Salience Network: A Central Pathway in Psychiatric Disease and Treatment. Front Syst Neurosci. 2016;10:104. doi:10.3389/fnsys.2016.00104

69. Tsuchiyagaito A, Misaki M, Cochran G, Philip NS, Paulus MP, Guinjoan SM. Thalamo-cortical circuits associated with trait- and state-repetitive negative thinking in major depressive disorder. J Psychiatr Res. 2023;168:184–192. doi:10.1016/j.jpsychires.2023.10.058

70. Admon R, Nickerson LD, Dillon DG, et al. Dissociable cortico-striatal connectivity abnormalities in major depression in response to monetary gains and penalties. Psychol Med. 2015;45(1):121–131. doi:10.1017/S0033291714001123

71. Schiller CE, Minkel J, Smoski MJ, Dichter GS. Remitted Major Depression is Characterized by Reduced Prefrontal Cortex Reactivity to Reward Loss. J Affect Disord. 2013;151(2):756–762. doi:10.1016/j.jad.2013.06.016

72. Koenigs M, Grafman J. The functional neuroanatomy of depression: Distinct roles for ventromedial and dorsolateral prefrontal cortex. Behav Brain Res. 2009;201(2):239–243. doi:10.1016/j.bbr.2009.03.004

73. Rogers MA, Kasai K, Koji M, et al. Executive and prefrontal dysfunction in unipolar depression: a review of neuropsychological and imaging evidence. Neurosci Res. 2004;50(1):1–11. doi:10.1016/j.neures.2004.05.003

74. Neumann WJ, Steiner LA, Milosevic L. Neurophysiological mechanisms of deep brain stimulation across spatiotemporal resolutions. Brain. 2023;146(11):4456–4468. doi:10.1093/brain/awad239

75. Harmsen IE, Wolff Fernandes F, Krauss JK, Lozano AM. Where Are We with Deep Brain Stimulation? A Review of Scientific Publications and Ongoing Research. Stereotact Funct Neurosurg. 2022;100(3):184–197. doi:10.1159/000521372

76. Provenza NR, Reddy S, Allam AK, et al. Disruption of neural periodicity predicts clinical response after deep brain stimulation for obsessive-compulsive disorder. Nat Med. 2024;30(10):3004–3014. doi:10.1038/s41591-024-03125-0

77. Safadi Z, Grisot G, Jbabdi S, et al. Functional Segmentation of the Anterior Limb of the Internal Capsule: Linking White Matter Abnormalities to Specific Connections. J Neurosci. 2018;38(8):2106–2117. doi:10.1523/JNEUROSCI.2335-17.2017

78. Sretavan K, Braun H, Liu Z, et al. A Reproducible Pipeline for Parcellation of the Anterior Limb of the Internal Capsule. Biol Psychiatry Cogn Neurosci Neuroimaging. 2024;9(12):1249–1261. doi:10.1016/j.bpsc.2024.07.008

79. Haber SN, Kim KS, Mailly P, Calzavara R. Reward-Related Cortical Inputs Define a Large Striatal Region in Primates That Interface with Associative Cortical Connections, Providing a Substrate for Incentive-Based Learning. J Neurosci. 2006;26(32):8368–8376. doi:10.1523/JNEUROSCI.0271-06.2006

80. Joyce MKP, Barbas H. Cortical Connections Position Primate Area 25 as a Keystone for Interoception, Emotion, and Memory. J Neurosci. 2018;38(7):1677–1698. doi:10.1523/JNEUROSCI.2363-17.2017

81. Nagao KJ, Perera T, Petoe M, et al. Automating Subthalamic Deep Brain Stimulation Programming with Evoked Resonant Neural Activity in Parkinson’s Disease. Mov Disord. n/a(n/a). doi:10.1002/mds.70318

82. Strelow JN, Dembek TA, Baldermann JC, et al. Local Field Potential-Guided Contact Selection Using Chronically Implanted Sensing Devices for Deep Brain Stimulation in Parkinson’s Disease. Brain Sci. 2022;12(12):1726. doi:10.3390/brainsci12121726

83. Spooner RK, Bahners BH, Schnitzler A, Florin E. DBS-evoked cortical responses index optimal contact orientations and motor outcomes in Parkinson’s disease. NPJ Park Dis. 2023;9:37. doi:10.1038/s41531-023-00474-4

84. Muller M, van Leeuwen MFC, Hoffmann CF, et al. From subthalamic local field potentials to the selection of chronic deep brain stimulation contacts in Parkinson’s disease - A systematic review. Brain Stimulat. 2025;18(5):1499–1510. doi:10.1016/j.brs.2025.08.004

85. Van Bogaert T, Figee M, Kopell BH, et al. Electrophysiological Biomarkers Reflect Target Engagement and Response Using Deep Brain Stimulation for Obsessive-Compulsive Disorder. Biol Psychiatry Glob Open Sci. 2025;6(1):100609. doi:10.1016/j.bpsgos.2025.100609

86. Kuncel AM, Grill WM. Selection of stimulus parameters for deep brain stimulation. Clin Neurophysiol. 2004;115(11):2431–2441. doi:10.1016/j.clinph.2004.05.031

87. Dickey CW, Hassan U, Kawasaki H, et al. Transcranial magnetic stimulation to the dorsolateral prefrontal cortex modulates single-neuron activity in humans. *bioRxiv*. Preprint posted online March 18, 2026:2026.03.15.711839. doi:10.64898/2026.03.15.711839

88. Miller KJ, Müller KR, Hermes D. Basis profile curve identification to understand electrical stimulation effects in human brain networks. PLOS Comput Biol. 2021;17(9):e1008710. doi:10.1371/journal.pcbi.1008710

89. Gregg NM, Ojeda Valencia G, Pridalova T, et al. Thalamic Stimulation Induced Changes in Network Connectivity and Excitability in Epilepsy. Ann Neurol. 2026;99(3):748–760. doi:10.1002/ana.78087

90. Opri E, Isbaine F, Borgheai SB, et al. Deep brain stimulation-induced local evoked potentials outperform spectral features in spatial and clinical STN mapping. J Neural Eng. 2025;22(4):046055. doi:10.1088/1741-2552/adf99f

91. Mohan UR, Watrous AJ, Miller JF, et al. The effects of direct brain stimulation in humans depend on frequency, amplitude, and white-matter proximity. Brain Stimulat. 2020;13(5):1183–1195. doi:10.1016/j.brs.2020.05.009

92. Sisterson ND, Carlson AA, Rutishauser U, et al. Electrocorticography During Deep Brain Stimulation Surgery: Safety Experience From 4 Centers Within the National Institute of Neurological Disorders and Stroke Research Opportunities in Human Consortium. Neurosurgery. 2021;88(5):E420–E426. doi:10.1093/neuros/nyaa592

93. Ljulj H, Lehner K, Wyse-Sookoo K, et al. Electrocorticography During Deep Brain Stimulation Surgery for Movement Disorders: Single-Center Experience. Brain Sci. 2026;16(6):561. doi:10.3390/brainsci16060561

94. Horn A, Fox MD. Opportunities of connectomic neuromodulation. NeuroImage. 2020;221:117180. doi:10.1016/j.neuroimage.2020.117180

95. Siddiqi SH, Fox MD. Targeting Symptom-Specific Networks With Transcranial Magnetic Stimulation. Biol Psychiatry. 2024;95(6):502–509. doi:10.1016/j.biopsych.2023.11.011

96. Mattar LS, Shah S, Chamakura LS, et al. Neural signatures and personalized neuromodulation in a subject experiencing context-dependent inhibitory control deficits. NeuroImage. 2025;321:121507. doi:10.1016/j.neuroimage.2025.121507

97. Bahners BH, Goede LL, Meyer GM, et al. Evoked response signatures explain deep brain stimulation outcomes. medRxiv. Preprint posted online October 6, 2024:2024.10.04.24314308. doi:10.1101/2024.10.04.24314308

98. Personalizing Brain Stimulation for Psychiatric Disorders: From Circuits to Closed-Loop Control | American Journal of Psychiatry. Accessed June 23, 2026. https://psychiatryonline.org/doi/10.1176/appi.ajp.20251327

99. Fitoz EC, Alagapan S, Cha J, et al. Common Electrophysiology Biomarkers Collected at Home Robustly Track Depression Recovery With Deep Brain Stimulation. medRxiv. Published online April 20, 2026:2026.04.13.26350107. doi:10.64898/2026.04.13.26350107

