## supplementary Material for "Stimulation-evoked orbitofrontal activity as a biomarker for DBS personalization in depression"

**Best DBS configurations yield stronger and more prevalent CCEPs in OFC: Full models**

Mixed-effects models to evaluate differences between CCEP prominence according to DBS configurations (“best” versus “other”; “other” is used as baseline level and thus is incorporated in the intercept coefficient; patients are included as random effects). Model 1 is the simplest, starting model, and more complex models (i.e., with additional coefficients) are tested progressively: with Model 2 we tested for spatial specificity by including a term identifying the anatomical locations of the contacts (OFC, VPFC, DPFC, DACC, LT, MTL and WM; WM used as baseline level) and evaluating the interaction between each anatomical location and configuration type; with Model 3 we additionally tested if the effects were dependent on the DBS lead position (SCC and VC/VS; VC/VS used as baseline level). For each model, the effects of interest are those displaying a significant interaction with best config, highlighted in gray. Hierarchical nested model comparisons using Likelihood Ratio Test were performed to test whether the additional model coefficients significantly improved the model fit.

Model 1 versus Model 2: χ^2^(12)=847.72, p<0.001

Model 2 versus Model 3: χ^2^(14)=845.15, p<0.001

**Model 1: CCEP ~ best config + (1 | subject)**

|  | **CCEP prominence** | | |
| --- | --- | --- | --- |
| *Predictors* | *Estimates* | *CI* | *p* |
| (Intercept) | 4.77 | 2.62 – 6.93 | **<0.001** |
| best config [1] | 2.46 | 1.78 – 3.13 | **<0.001** |
| **Random Effects** | | | |
| σ^2^ | 276.47 | | |
| τ_00_ _subject_ | 9.23 | | |
| ICC | 0.03 | | |
| N _subject_ | 8 | | |
| Observations | 10303 | | |
| Marginal R^2^ / Conditional R^2^ | 0.005 / 0.037 | | |

**Model 2: CCEP ~ best config * anatomy + (1 | subject)**

|  | **CCEP prominence** | | |
| --- | --- | --- | --- |
| *Predictors* | *Estimates* | *CI* | *p* |
| (Intercept) | 4.75 | 2.31 – 7.20 | **<0.001** |
| best config [1] | 0.02 | -2.04 – 2.08 | 0.987 |
| anat [dACC] | -3.12 | -5.38 – -0.85 | **0.007** |
| anat [DPFC] | -3.24 | -4.69 – -1.80 | **<0.001** |
| anat [LT] | -2.65 | -4.43 – -0.87 | **0.004** |
| anat [MTL] | -2.86 | -4.80 – -0.91 | **0.004** |
| anat [OFC] | 4.79 | 3.33 – 6.26 | **<0.001** |
| anat [VPFC] | 1.61 | -0.46 – 3.67 | 0.128 |
| best config [1] × anat [dACC] | 0.17 | -3.50 – 3.84 | 0.926 |
| best config [1] × anat [DPFC] | 0.77 | -1.57 – 3.11 | 0.517 |
| best config [1] × anat [LT] | 0.39 | -2.50 – 3.28 | 0.790 |
| best config [1] × anat [MTL] | 1.12 | -2.00 – 4.24 | 0.481 |
| best config [1] × anat [OFC] | 6.54 | 4.19 – 8.90 | **<0.001** |
| best config [1] × anat [VPFC] | 0.72 | -2.54 – 3.97 | 0.666 |
| **Random Effects** | | | |
| σ^2^ | 254.62 | | |
| τ_00_ _subject_ | 9.04 | | |
| ICC | 0.03 | | |
| N _subject_ | 8 | | |
| Observations | 10303 | | |
| Marginal R^2^ / Conditional R^2^ | 0.082 / 0.114 | | |

**Model 3: CCEP ~ best config * anatomy * DBSlead + (1 | subject)**

|  | **CCEP prominence** | | |
| --- | --- | --- | --- |
| *Predictors* | *Estimates* | *CI* | *p* |
| (Intercept) | 4.96 | 2.33 – 7.60 | **<0.001** |
| best config [1] | -1.83 | -4.58 – 0.92 | 0.193 |
| anat [dACC] | -4.22 | -7.25 – -1.19 | **0.006** |
| anat [DPFC] | -1.88 | -3.75 – -0.01 | **0.049** |
| anat [LT] | -1.49 | -3.81 – 0.83 | 0.208 |
| anat [MTL] | -2.62 | -5.15 – -0.10 | **0.041** |
| anat [OFC] | -0.44 | -2.34 – 1.47 | 0.652 |
| anat [VPFC] | 5.62 | 2.99 – 8.24 | **<0.001** |
| DBSlead [SCC] | -0.62 | -3.04 – 1.80 | 0.614 |
| best config [1] × anat [dACC] | 1.29 | -3.71 – 6.29 | 0.614 |
| best config [1] × anat [DPFC] | 0.03 | -3.10 – 3.16 | 0.985 |
| best config [1] × anat [LT] | 0.09 | -3.81 – 3.99 | 0.964 |
| best config [1] × anat [MTL] | 1.63 | -2.57 – 5.83 | 0.448 |
| best config [1] × anat [OFC] | 2.41 | -0.75 – 5.57 | 0.135 |
| best config [1] × anat [VPFC] | -0.23 | -4.61 – 4.15 | 0.917 |
| best config [1] × DBSlead [SCC] | 3.71 | -0.25 – 7.67 | 0.066 |
| anat [dACC] × DBSlead [SCC] | 2.40 | -1.92 – 6.73 | 0.276 |
| anat [DPFC] × DBSlead [SCC] | -2.86 | -5.61 – -0.11 | **0.042** |
| anat [LT] × DBSlead [SCC] | -2.32 | -5.73 – 1.09 | 0.182 |
| anat [MTL] × DBSlead [SCC] | -0.35 | -4.07 – 3.37 | 0.854 |
| anat [OFC] × DBSlead [SCC] | 11.00 | 8.23 – 13.77 | **<0.001** |
| anat [VPFC] × DBSlead [SCC] | -9.41 | -13.37 – -5.46 | **<0.001** |
| (best config [1] × anat [dACC]) × DBSlead [SCC] | -2.41 | -9.46 – 4.63 | 0.502 |
| (best config [1] × anat [DPFC]) × DBSlead [SCC] | 1.60 | -2.90 – 6.09 | 0.486 |
| (best config [1] × anat [LT]) × DBSlead [SCC] | 0.66 | -4.90 – 6.21 | 0.817 |
| (best config [1] × anat [MTL]) × DBSlead [SCC] | -1.11 | -7.11 – 4.89 | 0.717 |
| (best config [1] × anat [OFC]) × DBSlead [SCC] | 6.88 | 2.36 – 11.39 | **0.003** |
| (best config [1] × anat [VPFC]) × DBSlead [SCC] | 3.60 | -2.67 – 9.88 | 0.260 |
| **Random Effects** | | | |
| σ^2^ | 234.56 | | |
| τ_00_ _subject_ | 8.71 | | |
| ICC | 0.04 | | |
| N _subject_ | 8 | | |
| Observations | 10303 | | |
| Marginal R^2^ / Conditional R^2^ | 0.153 / 0.183 | | |

To better understand the three way interaction between best configuration, anatomical location of the CCEPs and DBS lead stimulated, we performed two separate mixed-effects models analogous to Model 2 for each DBS lead type.

**Model 2 SCC only: CCEP ~ best config * anatomy + (1 | subject)**

|  | **CCEP prominence** | | |
| --- | --- | --- | --- |
| *Predictors* | *Estimates* | *CI* | *p* |
| (Intercept) | 5.65 | 2.27 – 9.03 | **0.001** |
| best config [1] | 0.98 | -2.53 – 4.49 | 0.585 |
| anat [dACC] | -1.20 | -5.03 – 2.63 | 0.539 |
| anat [DPFC] | -4.43 | -6.94 – -1.91 | **0.001** |
| anat [LT] | -3.55 | -6.65 – -0.45 | **0.025** |
| anat [MTL] | -2.40 | -5.79 – 0.98 | 0.164 |
| anat [OFC] | 11.06 | 8.52 – 13.60 | **<0.001** |
| anat [VPFC] | -3.00 | -6.70 – 0.70 | 0.112 |
| best config [1] × anat [dACC] | -1.73 | -7.84 – 4.37 | 0.578 |
| best config [1] × anat [DPFC] | 0.95 | -3.02 – 4.92 | 0.639 |
| best config [1] × anat [LT] | 0.19 | -4.67 – 5.06 | 0.938 |
| best config [1] × anat [MTL] | -0.81 | -6.09 – 4.46 | 0.762 |
| best config [1] × anat [OFC] | 8.40 | 4.42 – 12.38 | **<0.001** |
| best config [1] × anat [VPFC] | 1.70 | -3.84 – 7.24 | 0.547 |
| **Random Effects** | | | |
| σ^2^ | 354.35 | | |
| τ_00_ _subject_ | 12.89 | | |
| ICC | 0.04 | | |
| N _subject_ | 8 | | |
| Observations | 5024 | | |
| Marginal R^2^ / Conditional R^2^ | 0.169 / 0.198 | | |

**Model 2 VC/VS only: CCEP ~ best config * anatomy + (1 | subject)**

|  | **CCEP prominence** | | |
| --- | --- | --- | --- |
| *Predictors* | *Estimates* | *CI* | *p* |
| (Intercept) | 4.57 | 1.79 – 7.35 | **0.001** |
| best config [1] | -1.62 | -3.54 – 0.29 | 0.096 |
| anat [dACC] | -4.64 | -6.74 – -2.54 | **<0.001** |
| anat [DPFC] | -1.94 | -3.24 – -0.64 | **0.003** |
| anat [LT] | -1.54 | -3.15 – 0.07 | 0.061 |
| anat [MTL] | -2.64 | -4.40 – -0.89 | **0.003** |
| anat [OFC] | -0.60 | -1.93 – 0.73 | 0.377 |
| anat [VPFC] | 5.61 | 3.78 – 7.44 | **<0.001** |
| best config [1] × anat [dACC] | 1.45 | -2.01 – 4.91 | 0.412 |
| best config [1] × anat [DPFC] | 0.15 | -2.02 – 2.31 | 0.893 |
| best config [1] × anat [LT] | 0.15 | -2.55 – 2.85 | 0.913 |
| best config [1] × anat [MTL] | 1.72 | -1.19 – 4.62 | 0.246 |
| best config [1] × anat [OFC] | 2.52 | 0.33 – 4.70 | **0.024** |
| best config [1] × anat [VPFC] | -0.33 | -3.36 – 2.70 | 0.832 |
| **Random Effects** | | | |
| σ^2^ | 112.13 | | |
| τ_00_ _subject_ | 13.22 | | |
| ICC | 0.11 | | |
| N _subject_ | 8 | | |
| Observations | 5279 | | |
| Marginal R^2^ / Conditional R^2^ | 0.034 / 0.136 | | |

**Higher odds of prefrontal CCEPs for best SCC configurations: full models**

Mixed-effects models to evaluate differences between the odds of detecting a CCEP according to DBS configurations (“best” versus “other”; “other” is used as baseline level and thus is incorporated in the intercept coefficient; patients are included as random effects). With Model-Odds 1 we tested for spatial specificity by including a term identifying the anatomical locations of the contacts (OFC, VPFC, DPFC, DACC, LT, MTL and WM; WM used as baseline level) and evaluating the interaction between each anatomical location and configuration type. We then ran the same model for SCC and VC/VS separately. The effects of interest are those displaying a significant interaction with best config, highlighted in gray.

**Model-Odds 1: CCEP detected ~ best config * anatomy + (1 | subject)**

|  | **CCEP detection** | | |
| --- | --- | --- | --- |
| *Predictors* | *Incidence Rate Ratios* | *CI* | *p* |
| (Intercept) | 0.19 | 0.12 – 0.29 | **<0.001** |
| best config [1] | 0.89 | 0.66 – 1.21 | 0.452 |
| anat [dACC] | 0.50 | 0.34 – 0.72 | **<0.001** |
| anat [DPFC] | 0.48 | 0.38 – 0.60 | **<0.001** |
| anat [LT] | 0.65 | 0.50 – 0.85 | **0.002** |
| anat [MTL] | 0.65 | 0.48 – 0.86 | **0.003** |
| anat [OFC] | 1.34 | 1.10 – 1.63 | **0.004** |
| anat [VPFC] | 0.80 | 0.59 – 1.08 | 0.140 |
| best config [1] × anat [dACC] | 1.12 | 0.59 – 2.11 | 0.732 |
| best config [1] × anat [DPFC] | 1.34 | 0.93 – 1.93 | 0.117 |
| best config [1] × anat [LT] | 1.19 | 0.76 – 1.85 | 0.452 |
| best config [1] × anat [MTL] | 1.55 | 0.98 – 2.45 | 0.059 |
| best config [1] × anat [OFC] | 1.46 | 1.05 – 2.02 | **0.023** |
| best config [1] × anat [VPFC] | 1.42 | 0.89 – 2.28 | 0.143 |
| **Random Effects** | | | |
| σ^2^ | 1.78 | | |
| τ_00_ _subject_ | 0.34 | | |
| ICC | 0.16 | | |
| N _subject_ | 8 | | |
| Observations | 10303 | | |
| Marginal R^2^ / Conditional R^2^ | 0.090 / 0.234 | | |

**Model-Odds 1 SCC only: CCEP detected ~ best config * anatomy + (1 | subject)**

|  | **CCEP detection** | | |
| --- | --- | --- | --- |
| *Predictors* | *Incidence Rate Ratios* | *CI* | *p* |
| (Intercept) | 0.25 | 0.17 – 0.38 | **<0.001** |
| best config [1] | 0.96 | 0.65 – 1.42 | 0.854 |
| anat [dACC] | 0.54 | 0.32 – 0.89 | **0.015** |
| anat [DPFC] | 0.14 | 0.09 – 0.21 | **<0.001** |
| anat [LT] | 0.38 | 0.24 – 0.59 | **<0.001** |
| anat [MTL] | 0.55 | 0.36 – 0.85 | **0.007** |
| anat [OFC] | 1.75 | 1.34 – 2.29 | **<0.001** |
| anat [VPFC] | 0.26 | 0.13 – 0.51 | **<0.001** |
| best config [1] × anat [dACC] | 1.18 | 0.55 – 2.56 | 0.671 |
| best config [1] × anat [DPFC] | 4.38 | 2.53 – 7.56 | **<0.001** |
| best config [1] × anat [LT] | 1.81 | 0.97 – 3.36 | 0.061 |
| best config [1] × anat [MTL] | 1.60 | 0.87 – 2.94 | 0.131 |
| best config [1] × anat [OFC] | 1.20 | 0.79 – 1.81 | 0.391 |
| best config [1] × anat [VPFC] | 2.66 | 1.16 – 6.08 | **0.020** |
| **Random Effects** | | | |
| σ^2^ | 1.52 | | |
| τ_00_ _subject_ | 0.23 | | |
| ICC | 0.13 | | |
| N _subject_ | 8 | | |
| Observations | 5024 | | |
| Marginal R^2^ / Conditional R^2^ | 0.331 / 0.419 | | |

**Model-Odds 1 VC/VS only: CCEP detected ~ best config * anatomy + (1 | subject)**

|  | **CCEP detection** | | |
| --- | --- | --- | --- |
| *Predictors* | *Incidence Rate Ratios* | *CI* | *p* |
| (Intercept) | 0.12 | 0.06 – 0.24 | **<0.001** |
| best config [1] | 0.64 | 0.39 – 1.07 | 0.089 |
| anat [dACC] | 0.46 | 0.26 – 0.81 | **0.007** |
| anat [DPFC] | 0.87 | 0.64 – 1.16 | 0.339 |
| anat [LT] | 0.98 | 0.69 – 1.40 | 0.924 |
| anat [MTL] | 0.79 | 0.53 – 1.18 | 0.252 |
| anat [OFC] | 0.93 | 0.69 – 1.25 | 0.634 |
| anat [VPFC] | 1.44 | 1.00 – 2.09 | 0.052 |
| best config [1] × anat [dACC] | 0.80 | 0.24 – 2.68 | 0.712 |
| best config [1] × anat [DPFC] | 0.78 | 0.43 – 1.40 | 0.398 |
| best config [1] × anat [LT] | 0.91 | 0.45 – 1.84 | 0.793 |
| best config [1] × anat [MTL] | 1.43 | 0.69 – 2.96 | 0.335 |
| best config [1] × anat [OFC] | 1.49 | 0.85 – 2.61 | 0.159 |
| best config [1] × anat [VPFC] | 1.44 | 0.72 – 2.87 | 0.302 |
| **Random Effects** | | | |
| σ^2^ | 2.00 | | |
| τ_00_ _subject_ | 0.97 | | |
| ICC | 0.33 | | |
| N _subject_ | 8 | | |
| Observations | 5279 | | |
| Marginal R^2^ / Conditional R^2^ | 0.038 / 0.352 | | |

**Supplementary analysis: Similar effects for medial and lateral OFC**

As a control analysis, we separated OFC into the medial (MOFC; n=1370) and lateral (LOFC; n=1791) subdivisions based on the Atlas labels and re-run the main analysis to test whether the effects were consistent between the two subdivisions.

Both MOFC and LOFC displayed a significant interaction with best configuration, resembling the same effect obtained when considering OFC as a whole, This validates our decision to not consider them separately. This result also confirms that monitoring OFC activity in response to DBS would not require high-precision anatomical data, as the CCEP modulation for best configuration can be measured across medial and lateral OFC.

**CCEP ~ best config * anatomy + (1 | subject)**

|  | **CCEP prominence** | | |
| --- | --- | --- | --- |
| *Predictors* | *Estimates* | *CI* | *p* |
| (Intercept) | 4.76 | 2.31 – 7.21 | **<0.001** |
| best config [1] | 0.02 | -2.04 – 2.07 | 0.988 |
| anat [dACC] | -3.12 | -5.39 – -0.86 | **0.007** |
| anat [DPFC] | -3.25 | -4.69 – -1.80 | **<0.001** |
| anat [LOFC] | 4.11 | 2.51 – 5.70 | **<0.001** |
| anat [LT] | -2.65 | -4.43 – -0.87 | **0.004** |
| anat [MOFC] | 5.68 | 4.00 – 7.36 | **<0.001** |
| anat [MTL] | -2.86 | -4.80 – -0.92 | **0.004** |
| anat [VPFC] | 1.60 | -0.47 – 3.66 | 0.130 |
| best config [1] × anat [dACC] | 0.17 | -3.49 – 3.84 | 0.926 |
| best config [1] × anat [DPFC] | 0.77 | -1.56 – 3.11 | 0.516 |
| best config [1] × anat [LOFC] | 6.21 | 3.66 – 8.77 | **<0.001** |
| best config [1] × anat [LT] | 0.39 | -2.49 – 3.28 | 0.789 |
| best config [1] × anat [MOFC] | 6.98 | 4.30 – 9.67 | **<0.001** |
| best config [1] × anat [MTL] | 1.12 | -1.99 – 4.24 | 0.480 |
| best config [1] × anat [VPFC] | 0.72 | -2.53 – 3.97 | 0.665 |
| **Random Effects** | | | |
| σ^2^ | 254.34 | | |
| τ_00_ _subject_ | 9.02 | | |
| ICC | 0.03 | | |
| N _subject_ | 8 | | |
| Observations | 10303 | | |
| Marginal R^2^ / Conditional R^2^ | 0.083 / 0.115 | | |
